# Understanding how spatial interactions of built environment features shape substance-use risk among youth in a rapidly urbanizing Nigerian city

**DOI:** 10.64898/2026.08.14.26360469

**Authors:** Afolabi Oyapero, Idowu Afeez Adedoyin, Oyapero Oyejoke, Onuche Victor Oma-Ojo, Adetona Ifeoluwa Olamide

## Abstract

**Background:** Adolescent and youth substance use is an important public health concern in rapidly urbanizing low- and middle-income countries; however, evidence on how social networks and community substance-use environments jointly shape recent use remains limited, particularly in African megacities.

**Methods:** We conducted a cross-sectional, community-based, convergent mixed-methods study of adolescents and young adults aged 12–24 years in the Yaba Local Council Development Area, Lagos, Nigeria. Quantitative data were collected using a structured questionnaire adapted from established, international survey instruments. The primary outcome was self-reported substance use within the past 30 days. Key exposures included a Social Exposure Score incorporating substance use among friends and family members, membership in a substance-using peer group, and perceived easy community availability of substances. Multivariable logistic regression was used to examine factors associated with past-30-day substance use, followed by an interaction model to assess whether perceived availability modified the association between social exposure and recent use. Open-ended responses on community approaches to reducing substance use were thematically analyzed and integrated with quantitative findings through a joint display.

**Results:** Among 285 participants (mean age 18.9 years; 52.6% male), 66 (23.2%) reported past-30-day substance use and 84 (29.5%) reported lifetime polysubstance use. A Higher Social Exposure Score was associated with increased odds of past-30-day substance use (adjusted odds ratio [aOR]=2.18; 95% CI: 1.59–2.99; p<0.001), while perceived easy community availability was independently associated with recent use (aOR=3.22; 95% CI: 1.42–7.30; p=0.005). The interaction between social exposure and perceived availability was statistically significant (aOR=1.51; 95% CI: 1.01–2.27; p=0.047), indicating that the association between social exposure and recent use varied according to perceived availability. The marginal effect of a one-unit increase in the Social Exposure Score on the predicted probability of past-30-day use was +0.08 when easy availability was not reported and +0.18 when it was reported. Among lifetime substance users, past-30-day use was more common among polysubstance users than single substance users (50.0% vs. 22.0%; χ²[1]=16.51; p<0.001). Qualitative findings identified supply side law enforcement (43.5%), population awareness campaigns (24.6%), regulatory and legislative control (16.1%), enhanced parental supervision (14.7%), and economic and youth empowerment (13.3%) as prominent community-proposed solutions. Integrated analysis demonstrated complementarity between the quantitatively identified social and environmental correlates and community-proposed intervention priorities.

**Conclusions:** In this urban Nigerian setting, past-30-day substance use was independently associated with social exposure and perceived community availability, with evidence that the association between social exposure and recent use varied according to perceived availability. The findings support multilevel prevention approaches that address environmental access alongside peer, family, community, and broader socioeconomic influences.

## Introduction

Adolescent substance use is a major global public health concern with important implications for neurodevelopment, mental health, educational attainment, and long-term socioeconomic outcomes.¹ A growing body of evidence indicates that substance use disorders and related harms contribute substantially to morbidity and disability during adolescence and young adulthood,² with initiation commonly occurring during these developmental periods.³ Although prevalence varies across settings, systematic reviews from sub-Saharan Africa indicate substantial levels of lifetime and current substance use among young people, with alcohol frequently reported alongside cannabis, stimulants, and other substances. Emerging evidence also suggests an increasing use of both licit and illicit substances among youth in low- and middle-income countries (LMICs), including sub-Saharan Africa, reflecting changing social environments, urbanization, and evolving patterns of access.

In Nigeria, Africa’s most populous country, rapid urbanization, socioeconomic precarity, and fragmented regulatory systems combine to create a complex risk environment for adolescents and young adults (AYAs). Community-based studies from Lagos have reported notable lifetime and recent substance use among secondary school students, including alcohol and non-medical opioid use such as tramadol, indicating both established and emerging patterns of use among urban youth.□ ⁻¹□ Adolescent substance use is shaped by interconnected family, peer, and social contexts, with peer and familial substance use, childhood trauma, adverse experiences, and demographic and socioeconomic factors emerging as important correlates of adolescent substance use. Protective influences, including positive parenting, supportive school relationships, and clear family and community disapproval of substance use, have also been associated with lower levels of polysubstance use, underscoring the importance of familial and institutional environments.^11,12^ Despite a growing national evidence base, the ecological and social determinants of earlier initiation and polysubstance involvement remain insufficiently characterized in Nigerian urban settings. Understanding these dynamics is essential for developing interventions that are both effective and contextually appropriate to the local setting.

Historically, adolescent substance-use research has emphasized individual-level factors, such as impulsivity, internalizing symptoms, sensation seeking, and family history of substance use. While these approaches have clarified important psychological and genetic vulnerabilities, they often underrepresent the broader social and environmental conditions within which behavior takes place. Social-ecological frameworks provide a more comprehensive perspective by conceptualizing substance use as the product of interactions across multiple nested systems, from interpersonal relationships to community environments and policy contexts,^13^ wherein peer influence, family context, community norms, and structural availability are important determinants of substance-use behavior. Peer networks are among the most consistently documented social influences, with adolescents exposed to peers who use substances more likely to initiate and maintain use.¹ Meta-analytic evidence further shows that peer influence operates both directly and through perceived norms across multiple substance-use behaviors.¹□ In multinational studies and systematic reviews of African adolescent populations, exposure to peer substance use has similarly emerged as an important correlate of use.¹□ Familial contexts also contribute, with parenting practices influencing perceptions of harm and normative attitudes that may modify the risk of polysubstance involvement.¹ □ However, relatively few studies in African urban settings have integrated these interpersonal influences with broader community and environmental exposures within a single socioecological framework.

A substantial body of literature demonstrates that the physical and economic accessibility of substances within communities can shape youth consumption. In alcohol research, higher outlet density and easier access have been associated with increased consumption and earlier initiation of alcohol consumption among adolescents.

Epidemiological studies linking both perceived availability and objective outlet density with adolescent drinking patterns support the “availability hypothesis,” whereby greater opportunity structures facilitate use.¹ Reviews of geographic availability and adolescent consumption similarly indicate that easier access tends to correspond to higher levels of use across diverse settings.¹□ Emerging evidence suggests that comparable mechanisms may also operate for other substances, including tobacco.² In under-regulated urban LMIC environments such as many Nigerian cities, however, access may extend beyond formal outlets to include unlicensed pharmacies, informal vendors, and street-based sellers locally described as “drug hawkers.” Although such supply channels are frequently identified in qualitative and mixed-methods research, their empirical relationship with adolescent and youth substance use patterns remains insufficiently quantified.

A critical unresolved question is whether social and environmental determinants interact to increase the risk of substance use. Evidence suggests that permissive availability may amplify peer influence by strengthening normative reinforcement and reducing access barriers. However, data from urban African settings are limited. Another important gap concerns the translation of epidemiological findings into locally acceptable and feasible interventions. Much of the evidence guiding global prevention strategies originates from Western contexts and may not fully reflect Nigerian urban environments in terms of their culture, economy, or regulation. Community-engaged mixed-methods approaches have therefore been proposed as important for bridging this “know-do gap” by incorporating youth perspectives into evidence-based intervention designs. Effective prevention increasingly requires action across multiple ecological levels, including individual skills, family and school support, peer norms, and community availability. Reviews of adolescent substance-use risk and protective factors similarly indicate that multilevel, culturally adapted strategies may be more sustainable when informed by the experiences of affected youth.

This study aimed to deepen the understanding of substance-use behaviors among adolescents and young adults in urban Lagos, Nigeria, using a theory-informed mixed-methods approach. It quantified the prevalence and patterns of substance use, examined the associations of social exposure and perceived community availability with recent substance use within a social-ecological framework, and tested whether perceived availability modified the association between social exposure and recent use of substances. The quantitative findings were then integrated with youth-generated qualitative data to identify contextually grounded multilevel intervention priorities.

Collectively, these components were intended to inform prevention approaches that are epidemiologically grounded and responsive to local conditions.

## Materials and Methods

### Study Design and Setting

We conducted a cross-sectional, community-based convergent mixed-methods study in the Yaba Local Council Development Area (LCDA), Lagos State, Nigeria, between November 2024 and April 2025. Yaba is a densely populated urban area comprising communities with varied socioeconomic characteristics and several educational and commercial centers. A convergent mixed-methods design was used to collect quantitative and qualitative data during the same period. The two data strands were analyzed separately and subsequently integrated during interpretation to provide complementary insights into the patterns and determinants of substance use and the participants’ perspectives on potential community-level solutions.

### Population and Sampling Strategy

The study population comprised adolescents and young adults aged 12–24 years residing in the selected study areas within the Yaba LCDA. A multistage non-probability sampling strategy combining venue-based and peer-referral recruitment was used. Five study areas were purposively selected to capture variations in socioeconomic and environmental characteristics. Within these areas, participants were recruited from locations frequented by young people, including motor parks, market areas and educational institutions. Limited peer referral, with a maximum of two referrals per participant, was used to reach eligible young people who might not have been encountered at the selected recruitment venues.

Eligible participants were aged 12–24 years and resided in one of the selected study areas of the country. Individuals who were unable to provide informed consent or assent because of apparent cognitive impairment or acute intoxication at the time of recruitment were excluded from the study. Of the 365 eligible individuals invited to participate, 285 enrolled in the study, corresponding to a participation rate of 78.1%.

### Data Collection Instrument and Procedures

A structured questionnaire was developed by adapting items from established survey instruments, including the United Nations Office on Drugs and Crime (UNODC) drug-use survey resources and the World Health Organization Global School-based Student Health Survey (GSHS).^21,22^ The questionnaire comprised four domains: (1) sociodemographic and household characteristics; (2) substance-use patterns, including lifetime, past year, and past-30-day use across ten substance categories, age at initiation, and sequence of substance-use initiation; (3) social-ecological determinants, including peer and family substance use, perceived community availability, and sources of substances; and (4) perceived consequences of substance use and open-ended suggestions for reducing substance use in participants’ communities.

Data were collected through face-to-face interviews conducted by six trained research assistants with public health and community-based work backgrounds. The interviews lasted approximately 25 minutes and were conducted in private settings to protect confidentiality and ensure the safety of the interviewers. Interviews were not audio-recorded to enhance privacy and participant comfort while discussing sensitive behaviors. Quality control procedures included daily supervisory review of completed questionnaires and random back-checks of 10% of completed interviews.

### Measures

The primary outcome was past-30-day substance use, defined as the self-reported use of at least one of the following substances during the preceding 30 days: alcohol, tobacco, cannabis, tramadol, cocaine, solvents/inhalants, or other specified psychoactive substances. The outcome was coded dichotomously as any versus no past-30-day use.

The key independent variables included:

1. **Social Exposure Index:** A composite score derived by summing three binary indicators: having substance-using friends, having substance-using family members, and belonging to a peer group or clique in which substances were used. Scores ranged from 0 to 3, with higher scores indicating greater social exposure to substance use (Cronbach’s α=0.72).
2. **Perceived Community Availability:** Assessed using the question, “Are these substances easily available in your neighborhood?” and coded as yes/no.
3. **Polysubstance Use:** Defined as the lifetime use of two or more substance classes and coded dichotomously.
4. **Sociodemographic covariates included** age (<18 vs. ≥18 years), gender, educational attainment, parental education, family structure, living arrangement, and estimated monthly household income (< 50,000, □50,000–□100,000, and >□100,000).

### Ethical Considerations

This study was approved by the Health Research and Ethics Committee of Lagos State Health Service Commission. Multiple safeguards were implemented to protect the participants. All interviewers completed Good Clinical Practice training, participants were provided with study information in Yoruba and Pidgin English as appropriate, and referral pathways to local substance-use treatment services were established. Study data were de-identified immediately after collection using unique study identifiers and were stored on password-protected, encrypted devices. Written informed consent was obtained from participants who could legally provide consent. For participants younger than 16 years, written permission was obtained from a parent or legal guardian, and assent was subsequently obtained from the minor after the study purpose, procedures, potential risks, and benefits were explained in a language appropriate to their level of understanding. Participation was voluntary, and participants, and where applicable, their parents or guardians, were informed of their right to withdraw at any time without penalty or adverse consequences.

### Quantitative Analysis

Data were analyzed using IBM SPSS Statistics version 26.0 (IBM Corp., Armonk, NY, USA), with statistical significance set at *p*<0.05. Descriptive statistics were used to summarize the participants’ characteristics and study variables. Categorical variables were summarized using frequencies and percentages, and continuous variables were summarized using means and standard deviations or medians and interquartile ranges, as appropriate. Bivariate associations were examined using χ² tests for categorical variables and independent-samples *t*-tests or Mann–Whitney U tests for continuous variables, as appropriate. Sequential multivariable logistic regression models were constructed to examine the factors associated with past-30-day tobacco or nicotine use. Model 1 included sociodemographic covariates; Model 2 included the Social Exposure Index; and Model 3 included Perceived Community Availability. Model calibration was assessed using the Hosmer–Lemeshow goodness-of-fit test, and discrimination was evaluated using the area under the receiver operating characteristic curve (AUC).

Multicollinearity was assessed using variance inflation factors (VIFs), all of which were <2.5. To test the primary hypothesis that perceived community availability moderated the association between social exposure and past-30-day tobacco or nicotine use, an interaction model was fitted that included the main effects of the Social Exposure Index and Perceived Community Availability and their product term (Social Exposure Index × Perceived Community Availability), together with the prespecified covariates. Where evidence of interaction was observed, conditional effects were examined using model-based adjusted predicted probabilities across relevant values of the interacting variables and were graphically displayed to facilitate interpretation.

### Qualitative Analysis

Open-ended responses to the question, “How can substance abuse be reduced in your neighborhood?” were analyzed using reflexive thematic analysis. Two researchers independently coded the responses in NVivo 14 and iteratively developed and refined the consensus codebook. The initial coding was data-driven, and the resulting themes were mapped onto a social-ecological framework encompassing the individual, interpersonal, community, and structural levels. Intercoder agreement was assessed for 20% of the responses (Cohen’s κ=0.82), with coding discrepancies resolved through discussion and consensus. Participant feedback was obtained from five participants to assess the resonance and credibility of the researchers’ interpretation.

### Integration Methodology

The quantitative and qualitative findings were integrated using a joint display approach (6). Quantitative risk factors and qualitative solution themes were juxtaposed in a matrix to identify areas of confirmation, where findings from the two strands were concordant; complementarity, where the findings provided different but mutually informative insights; and divergence, where the findings differed and required further interpretation. Integration occurred during the interpretation phase after quantitative and qualitative analyses were conducted separately.

## Results

Figure 1 shows the Spatial Distribution of Past-30-Day Substance Use and Contextual Risk Factors Across the Five Geographical Study Areas in Yaba LCDA, Lagos State, Nigeria. Panel A depicts the geographical distribution of the five study areas—Onike, Makoko, Iwaya, Alagomeji, and Abule Oja—and selected urban and environmental features relevant to the study context, including major roads, railway infrastructure, markets, motor parks, educational institutions, and water bodies. Panel B presents the spatial distribution of selected substance use and contextual indicators across the five study areas, including past-30-day substance use, mean Social Exposure Scores, perceived easy community availability of substances, commonly reported substance sources, and participant-reported substance-use locations or hotspots. Past-30-day substance-use prevalence was 23.4% in Onike, 34.5% in Makoko, 23.6% in Iwaya, 19.7% in Alagomeji, and 12.8% in Abule Oja. Color gradients indicate variations in prevalence or mean values across study areas, while symbols identify selected contextual and environmental features. Geographical differences in past-30-day substance use were not statistically significant (Pearson’s χ²[4]=7.46, p=0.114).

**Figure 1:**
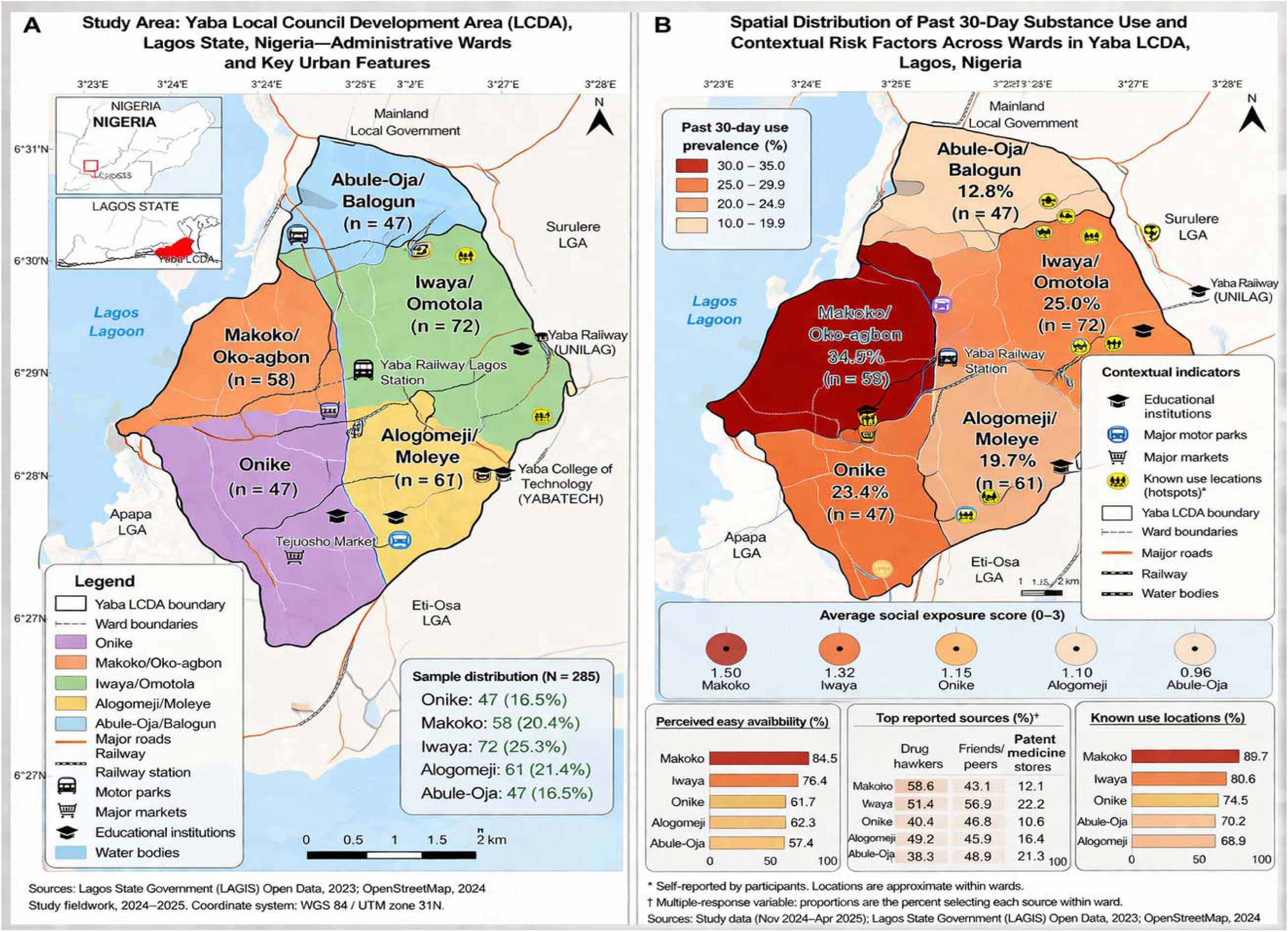
Spatial Distribution of Past 30-Day Substance Use and Contextual Risk Factors Across Wards in Yaba LCDA, Lagos, Nigeria, with Youth-Reported Hotspots and Environmental Exposures.

Table 1 presents demographic, socioeconomic, substance use, social exposure, community environment, and substance source characteristics for the total sample and separately for participants recruited from Onike, Makoko, Iwaya, Alagomeji, and Abule Oja. Categorical variables are presented as frequencies and percentages, while the Social Exposure Score is presented as the mean and standard deviation. Pearson’s χ² tests were used to examine differences in categorical characteristics across the five independent geographical study areas, and one-way analysis of variance was used to compare the mean Social Exposure Scores. Effect sizes are presented as Cramer’s V for categorical variables and η² for ANOVA. Polysubstance use was defined as the lifetime use of two or more substance classes. The Social Exposure Score ranged from 0 to 3 and comprised binary indicators of substance use among friends, substance use among family members, and membership in a substance-using peer group or clique.

**Table 1.** Sociodemographic, Substance-Use, and Contextual Characteristics by Geographical Study Area.

| Characteristic | Total (N=285) | Onike (n=47) | Makoko (n=58) | Iwaya (n=72) | Alagomeji (n=61) | Abule Oja (n=47) | Statistical test | p-value | Effect size |
| --- | --- | --- | --- | --- | --- | --- | --- | --- | --- |
| **Demographic characteristics** |  |  |  |  |  |  |  |  |  |
| Age ≥18 years | 132 (46.3) | 18 (38.3) | 28 (48.3) | 37 (51.4) | 27 (44.3) | 22 (46.8) | $\chi^2(4)=2.16$ | 0.707 | Cramer's V=0.087 |
| Male | 150 (52.6) | 22 (46.8) | 33 (56.9) | 41 (56.9) | 31 (50.8) | 23 (48.9) | $\chi^2(4)=1.94$ | 0.747 | Cramer's V=0.082 |
| **Socioeconomic characteristics** |  |  |  |  |  |  |  |  |  |
| Tertiary education | 65 (22.8) | 6 (12.8) | 8 (13.8) | 19 (26.4) | 18 (29.5) | 14 (29.8) | $\chi^2(4)=8.75$ | 0.068 | Cramer's V=0.175 |
| Parental tertiary education | 98 (34.4) | 11 (23.4) | 13 (22.4) | 31 (43.1) | 24 (39.3) | 19 (40.4) | $\chi^2(4)=10.02$ | **0.040** | Cramer's V=0.188 |
| Household income > ₦100,000 | 77 (27.0) | 9 (19.1) | 11 (19.0) | 23 (31.9) | 20 (32.8) | 14 (29.8) | $\chi^2(4)=5.48$ | 0.241 | Cramer's V=0.139 |
| **Substance-use characteristics** |  |  |  |  |  |  |  |  |  |
| Lifetime substance use | 193 (67.7) | 32 (68.1) | 45 (77.6) | 52 (72.2) | 39 (63.9) | 25 (53.2) | $\chi^2(4)=8.19$ | 0.085 | Cramer's V=0.170 |
| **Past-30-day substance use** | **66 (23.2)** | **11 (23.4)** | **20 (34.5)** | **17 (23.6)** | **12 (19.7)** | **6 (12.8)** | **$\chi^2(4)=7.46$** | **0.114** | **Cramer's V=0.162** |
| Lifetime polysubstance | 84 (29.5) | 13 (27.7) | 22 (37.9) | 25 (34.7) | 16 (26.2) | 8 (17.0) | $\chi^2(4)=6.84$ | 0.145 | Cramer's V=0.155 |
| use† |  |  |  |  |  |  |  |  |  |
| **Contextual characteristics** |  |  |  |  |  |  |  |  |  |
| Social Exposure Score, mean (SD)‡ | 1.23 (0.98) | 1.15 (0.94) | 1.50 (1.02) | 1.32 (0.97) | 1.10 (0.95) | 0.96 (0.92) | F(4,280)=2.69 | **0.032** | $\eta^2=0.037$ |
| Perceived easy availability | 198 (69.5) | 29 (61.7) | 49 (84.5) | 55 (76.4) | 38 (62.3) | 27 (57.4) | $\chi^2(4)=13.81$ | **0.008** | Cramer's V=0.220 |
| Known substance-use locations | 220 (77.2) | 35 (74.5) | 52 (89.7) | 58 (80.6) | 42 (68.9) | 33 (70.2) | $\chi^2(4)=9.49$ | **0.050** | Cramer's V=0.182 |
| **Reported substance sources§** |  |  |  |  |  |  |  |  |  |
| Drug hawkers | 138 (48.4) | 19 (40.4) | 34 (58.6) | 37 (51.4) | 30 (49.2) | 18 (38.3) | $\chi^2(4)=5.82$ | 0.213 | Cramer's V=0.143 |
| Friends/peers | 139 (48.8) | 22 (46.8) | 25 (43.1) | 41 (56.9) | 28 (45.9) | 23 (48.9) | $\chi^2(4)=2.94$ | 0.567 | Cramer's V=0.102 |
| Patent medicine stores | 48 (16.8) | 5 (10.6) | 7 (12.1) | 16 (22.2) | 10 (16.4) | 10 (21.3) | $\chi^2(4)=4.39$ | 0.356 | Cramer's V=0.124 |

**Notes:** Values are n (%) unless otherwise indicated. †Polysubstance use was defined as lifetime use of $\geq 2$ substance classes. ‡The Social Exposure Score ranged from 0 to 3 and comprised indicators of substance use among friends, family members, and membership in substance-using peer groups/cliques; values are presented as mean (SD) and compared using one-way ANOVA. §Participants could report more than one source of substances. Each source was treated as a separate binary participant-level variable and compared across the five independent geographical study areas using Pearson $\chi^2$ tests. Cramer's V is reported as the effect-size measure for categorical comparisons and $\eta^2$ for the ANOVA

Participants could report more than one source of substance; consequently, each substance source category was treated as a separate participant-level binary variable and compared across study areas using Pearson’s χ² tests. Past-30-day substance use was reported by 66 of the 285 participants (23.2%), with no statistically significant difference across geographical study areas (χ²[4]=7.46, p=0.114).

Table 2 describes the first substance used, lifetime single- and polysubstance-use patterns, commonly reported substance combinations, and recency of substance use among the 193 participants who reported lifetime substance use. Percentages were calculated using the lifetime-user subsample as the denominator, unless otherwise specified. Polysubstance use was defined as the lifetime use of at least two substance classes. Sixty-six lifetime users (34.2%) reported substance use during the preceding 30 days, whereas 127 (65.8%) reported no past-30-day use. Among participants with lifetime polysubstance use, 42 of 84 (50.0%) reported past-30-day use compared with 24 of 109 (22.0%) single substance users. The association between lifetime use patterns and past-30-day use was statistically significant (Pearson χ²[1]=16.51, p<0.001; φ=0.292). The corresponding unadjusted odds ratio was 3.54 (95% CI 1.90– 6.60), indicating higher odds of past-30-day substance use among polysubstance users than single substance users.

**Table 2.** Substance-Use Initiation, Use Patterns, Recency, and Association with Past-30-Day Use Among Lifetime Substance Users (n=193)

| Section | Characteristic | n | % of lifetime users | Past-30-day use, n | No past-30-day use, n | Additional information |
| --- | --- | --- | --- | --- | --- | --- |
| **First substance used** | Alcohol | 161 | 83.4 | — | — | Most frequently reported first substance |
|  | Tobacco | 23 | 11.9 | — | — | — |
|  | Cannabis | 8 | 4.1 | — | — | — |
|  | Other substance | 1 | 0.5 | — | — | Solvent/inhalant |
| **Substance-use pattern** | Single-substance use | 109 | 56.5 | 24 | 85 | Primarily alcohol: 101/109 (92.7%); past-30-day prevalence=22.0% |
|  | Polysubstance use (≥2 substance classes) | 84 | 43.5 | 42 | 42 | Past-30-day prevalence=50.0% |
|  | Alcohol + tobacco | 52 | 26.9 | — | — | Most frequently reported combination |
|  | Alcohol + cannabis | 24 | 12.4 | — | — |  |
|  | Three or more substance classes | 8 | 4.1 | — | — |  |
| **Recency of substance use** | No past-30-day use | 127 | 65.8 | — | — | 127/193 lifetime users |
|  | Past-30-day use | 66 | 34.2 | — | — | 66/193 lifetime users |
| **Association by lifetime use pattern** | Polysubstance use | 84 | 43.5 | 42 | 42 | 50.0% reported past-30-day use |
|  | Single-substance use | 109 | 56.5 | 24 | 85 | 22.0% reported past-30-day use |
|  | Total | 193 | 100.0 | 66 | 127 | 34.2% reported past-30-day use |

**Note:** Percentages in the “% of lifetime users” column use the 193 participants reporting lifetime substance use as the denominator. Polysubstance use was defined as lifetime use of at least two substance classes. For the association between lifetime use pattern and past-30-day substance use, **Pearson $\chi^2(1)=16.51$ , $p<0.001$ ; $\phi=0.292$ . The unadjusted OR was 3.54 (95% CI 1.90–6.60)**, indicating higher odds of past-30-day use among polysubstance users than among single-substance users.

Table 3 presents adjusted odds ratios (aORs), 95% confidence intervals (CIs), p-values, and variance inflation factors (VIFs) from the multivariable logistic regression examining factors associated with past-30-day substance use. Covariates included age, sex, educational attainment, parental educational attainment, household income, Social Exposure Score, Perceived Easy Availability, and fixed effects for the geographical study area, with Onike serving as the reference category. A one-unit increase in the Social Exposure Score was associated with higher adjusted odds of past-30-day substance use (aOR=2.18, 95% CI 1.59–2.99; p<0.001), while perceived easy availability was also positively associated with past-30-day use (aOR=3.22, 95% CI 1.42–7.30; p=0.005). Model calibration was assessed using the Hosmer–Lemeshow goodness-of-fit test (χ²[8]=6.84, p=0.554). Additional model indices included a log-likelihood of −113.21, Cox & Snell R² of 0.234, and Nagelkerke R² of 0.382, respectively. All VIFs were below 2.5, indicating no evidence of multicollinearity.

**Table 3.** Multivariable Logistic Regression of Factors Associated With Past-30-Day Substance Use.

| **Predictor** | **aOR** | **95% CI** | **p-value** | **VIF** |
| --- | --- | --- | --- | --- |
| Age ≥18 years | 1.08 | 0.54–2.17 | 0.821 | 1.12 |
| Male sex | 1.67 | 0.89–3.15 | 0.110 | 1.08 |
| Tertiary education | 0.61 | 0.25–1.48 | 0.274 | 1.42 |
| Parental tertiary education | 0.70 | 0.32–1.54 | 0.378 | 1.21 |
| Household income: medium | 0.93 | 0.38–2.26 | 0.871 | 1.31 |
| Household income: high | 0.62 | 0.26–1.49 | 0.285 | 1.38 |
| **Social Exposure Score, per one-unit increase** | **2.18** | **1.59–2.99** | **<0.001** | 1.24 |
| **Perceived easy availability** | **3.22** | **1.42–7.30** | **0.005** | 1.19 |
| Makoko vs Onike | 1.52 | 0.58–3.97 | 0.394 | 1.67 |
| Iwaya vs Onike | 1.08 | 0.43–2.71 | 0.867 | 1.81 |
| Alagomeji vs Onike | 0.78 | 0.29–2.10 | 0.624 | 1.74 |
| Abule Oja vs Onike | 0.50 | 0.16–1.60 | 0.244 | 1.70 |

**Model diagnostics:** Log-likelihood=−113.21; Hosmer–Lemeshow $\chi^2(8)=6.84$ , $p=0.554$ ; Cox & Snell $R^2=0.234$ ; Nagelkerke $R^2=0.382$ . All variance inflation factors were <2.5.

Table 4. Moderated Logistic Regression Examining the Interaction Between Social Exposure and Perceived Easy Availability in Relation to Past-30-Day Substance Use

**Table 4.** Moderated Logistic Regression Examining the Interaction Between Social Exposure and Perceived Easy Availability.

| Section | Model term / availability condition | Estimate | 95% CI | p-value |
| --- | --- | --- | --- | --- |
| **Adjusted Logistic Regression Model** | Social Exposure Score | **aOR 1.66** | **1.13–2.44** | **0.010** |
|  | Perceived easy availability | aOR 1.81 | 0.59–5.59 | 0.302 |
|  | **Social Exposure Score × Perceived Easy Availability** | **aOR 1.51** | **1.01–2.27** | **0.047** |
| **Conditional Marginal Effects** | Easy availability not reported | +0.08 | 0.02–0.14 | 0.012 |
|  | Easy availability reported | +0.18 | 0.12–0.24 | <0.001 |
|  | Difference in marginal effects | +0.10 | 0.00–0.20 | 0.047 |

The model was adjusted for age, sex, educational attainment, parental education, household income, and geographical study area. Conditional marginal effects represent the estimated change in the predicted probability of past-30-day substance use associated with a one-unit increase in the Social Exposure Score at each level of Perceived Easy Availability.

Table 4 presents the moderation analysis examining whether perceived community availability modified the association between the Social Exposure Score and past-30-day substance use. Adjusted odds ratios, 95% confidence intervals, and p-values are presented for the Social Exposure Score, Perceived Easy Availability, and their multiplicative interaction term. The interaction between the Score and perceived easy availability was statistically significant (aOR=1.51, 95% CI 1.01–2.27; p=0.047). The table additionally reports the conditional marginal effects derived from the adjusted predictions. A one-unit increase in the Social Exposure Score corresponded to an estimated 0.08 increase in the predicted probability of past-30-day substance use when easy community availability was not reported (95% CI 0.02–0.14; p=0.012) and a 0.18 increase when easy availability was reported (95% CI 0.12–0.24; p<0.001). The estimated difference between these marginal effects was 0.10 (95% CI 0.00–0.20; p=0.047). The model was adjusted for age, sex, educational attainment, parental education, household income, and geographical area.

Table 5 summarizes the themes identified from participants’ open-ended responses concerning how substance use could be reduced in their communities. Themes were organized according to their social-ecological level and presented as frequencies and percentages of the full sample. Supply side law enforcement was the most frequently identified intervention theme (124/285, 43.5%), followed by population awareness campaigns (70/285, 24.6%), regulatory and legislative control (46/285, 16.1%), enhanced parental supervision (42/285, 14.7%), economic and youth empowerment (38/285, 13.3%), and rehabilitation and treatment access (18/285, 6.3%). Fatalism or resistance to change was expressed by 17 participants (6.0%), while 67 responses (23.5%) were non-codable or insufficiently specific responses. Individual responses could be assigned to more than one thematic category; therefore, the themes were not mutually exclusive, and the percentages sum to more than 100%. Illustrative quotations were identified using respondent IDs.

**Table 5.** Thematic Analysis of Community-Proposed Approaches to Reducing Substance Use (N=285)

| Social-ecological level | Theme | Operational definition | n (%) |
| --- | --- | --- | --- |
| Structural | Economic and youth empowerment | Provision of employment, skills training, economic opportunities, and community amenities intended to address socioeconomic contributors to substance use | 38 (13.3) |
| Structural | Regulatory and legislative control | Laws and regulations addressing substance pricing, age restrictions, production, importation, or distribution | 46 (16.1) |
| Community | **Supply-side law enforcement** | Enforcement against sellers and suppliers, including arrests, raids, restrictions on sales, and confiscation of substances | **124 (43.5)** |
| Community | Population awareness campaigns | Mass-media campaigns, school-based education, religious messages, and other community awareness activities | 70 (24.6) |
| Interpersonal | Enhanced parental supervision | Increased parental monitoring, advice, guidance, and supervision of young people | 42 (14.7) |
| Individual/service level | Rehabilitation and treatment access | Counselling, treatment, rehabilitation services, and establishment or expansion of treatment facilities | 18 (6.3) |
| Barrier to change | Fatalism and resistance | Expressions that substance use could not be reduced or resistance to proposed interventions | 17 (6.0) |
| — | Non-codable/non-specific responses | Responses such as “no idea,” “nil,” irrelevant statements, empty responses, or responses insufficiently specific for thematic coding | 67 (23.5) |

**Note:** Percentages are based on the full sample (N=285). Participants could provide responses coded to more than one theme; therefore, categories were not mutually exclusive and percentages sum to more than 100%.

Table 6 integrates quantitative and qualitative findings by juxtaposing statistically identified substance-use correlates with the corresponding community-proposed intervention themes. Quantitative findings included the association between Perceived Easy Availability and past-30-day substance use (aOR=3.22, 95% CI 1.42–7.30; p=0.005), the association between Social Exposure Score and past-30-day use (aOR=2.18, 95% CI 1.59–2.99; p<0.001), the interaction between Social Exposure Score and Perceived Easy Availability (aOR=1.51, 95% CI 1.01–2.27; p=0.047), and the association between lifetime polysubstance use and past-30-day use (50.0% vs. 22.0%; χ²[1]=16.51, p<0.001; unadjusted OR=3.54, 95% CI 1.90–6.60). These findings were integrated with community-proposed themes, including supply side law enforcement, awareness campaigns, enhanced parental supervision, and rehabilitation and treatment access. The joint display distinguishes areas of confirmation, complementarity, expansion, and divergence between quantitative and qualitative strands. Integrated interpretations are intended to characterize the convergence between data sources and should not be interpreted as evidence of causality or demonstrated intervention effectiveness.

**Table 6.**
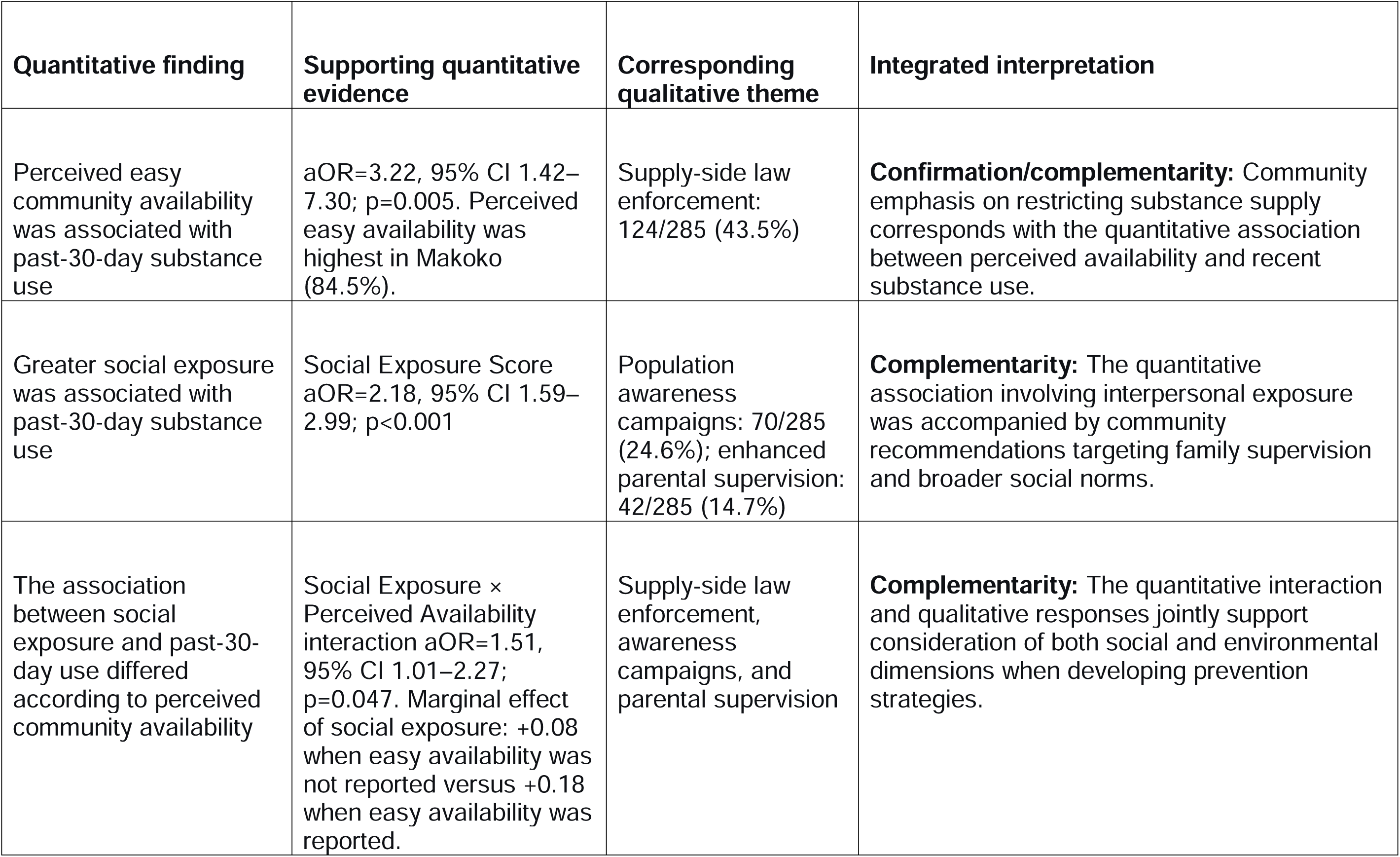
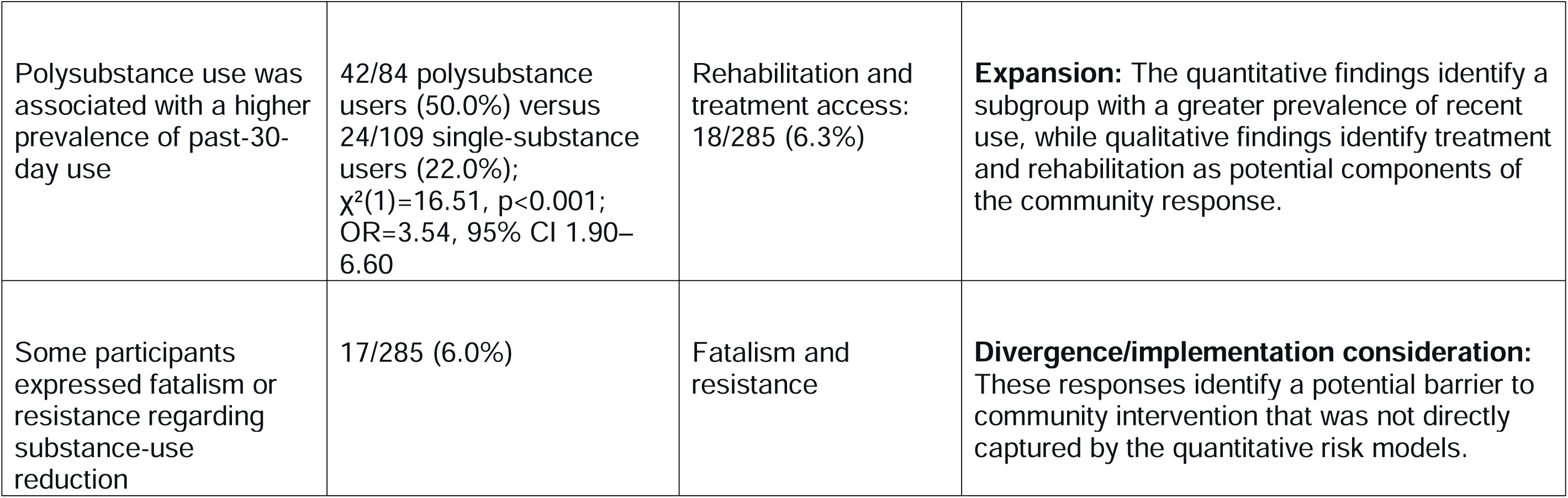
Integrative Joint Display Linking Quantitative Risk Indicators With Community-Proposed Intervention Themes.

| Quantitative finding | Supporting quantitative evidence | Corresponding qualitative theme | Integrated interpretation |
| --- | --- | --- | --- |
| Perceived easy community availability was associated with past-30-day substance use | aOR=3.22, 95% CI 1.42–7.30; p=0.005. Perceived easy availability was highest in Makoko (84.5%). | Supply-side law enforcement: 124/285 (43.5%) | **Confirmation/complementarity:** Community emphasis on restricting substance supply corresponds with the quantitative association between perceived availability and recent substance use. |
| Greater social exposure was associated with past-30-day substance use | Social Exposure Score aOR=2.18, 95% CI 1.59–2.99; p<0.001 | Population awareness campaigns: 70/285 (24.6%); enhanced parental supervision: 42/285 (14.7%) | **Complementarity:** The quantitative association involving interpersonal exposure was accompanied by community recommendations targeting family supervision and broader social norms. |
| The association between social exposure and past-30-day use differed according to perceived community availability | Social Exposure × Perceived Availability interaction aOR=1.51, 95% CI 1.01–2.27; p=0.047. Marginal effect of social exposure: +0.08 when easy availability was not reported versus +0.18 when easy availability was reported. | Supply-side law enforcement, awareness campaigns, and parental supervision | **Complementarity:** The quantitative interaction and qualitative responses jointly support consideration of both social and environmental dimensions when developing prevention strategies. |
| Polysubstance use was associated with a higher prevalence of past-30-day use | 42/84 polysubstance users (50.0%) versus 24/109 single-substance users (22.0%); $\chi^2(1)=16.51$, $p<0.001$; OR=3.54, 95% CI 1.90–6.60 | Rehabilitation and treatment access: 18/285 (6.3%) | **Expansion:** The quantitative findings identify a subgroup with a greater prevalence of recent use, while qualitative findings identify treatment and rehabilitation as potential components of the community response. |
| Some participants expressed fatalism or resistance regarding substance-use reduction | 17/285 (6.0%) | Fatalism and resistance | **Divergence/implementation consideration:** These responses identify a potential barrier to community intervention that was not directly captured by the quantitative risk models. |

Table 7 provides a consolidated summary of the principal descriptive, bivariate, multivariable, moderation, and qualitative findings. Lifetime substance use was reported by 193 of the 285 participants (67.7%), while 66 of the 285 (23.2%) reported past-30-day substance use. Among lifetime users, 66 of 193 (34.2%) reported past-30-day use, and 84 of 193 (43.5%) met the definition of lifetime polysubstance use. Half of polysubstance users (42/84, 50.0%) reported past-30-day substance use compared with 24 of 109 (22.0%) single-substance users (Pearson χ²[1]=16.51, p<0.001; φ=0.292; unadjusted OR=3.54, 95% CI 1.90–6.60). In the adjusted analyses, the Social Exposure (aOR=2.18, 95% CI 1.59–2.99; p<0.001) and Perceived Easy Availability (aOR=3.22, 95% CI 1.42–7.30; p=0.005) were associated with past-30-day substance use, and their interaction was statistically significant (aOR=1.51, 95% CI 1.01–2.27; p=0.047).

**Table 7.** Summary of Principal Quantitative and Mixed-Methods Findings.

| **Principal finding** | **Numerical evidence** | **Interpretation** |
| --- | --- | --- |
| Lifetime substance use was common | 193/285 (67.7%) | More than two-thirds of participants reported having ever used at least one substance. |
| Past-30-day substance use was reported by nearly one-quarter of participants | 66/285 (23.2%) | Approximately one in four participants reported substance use during the preceding 30 days. |
| Past-30-day use among lifetime users | 66/193 (34.2%) | Approximately one-third of participants with lifetime substance use reported use in the preceding 30 days. |
| Polysubstance use was common among lifetime users | 84/193 (43.5%); 84/285 (29.5% of full sample) | A substantial proportion of lifetime users reported use of at least two substance classes. |
| Past-30-day use was more common among polysubstance users | 42/84 (50.0%) vs 24/109 (22.0%); $\chi^2(1)=16.51$, $p<0.001$; $\phi=0.292$; OR=3.54, 95% CI 1.90–6.60 | Polysubstance users had substantially higher odds of recent use than single-substance users. |
| Greater social exposure was independently associated with recent substance use | aOR=2.18, 95% CI 1.59–2.99; $p<0.001$ | Each one-unit increase in the Social Exposure Score was associated with higher adjusted odds of past-30-day substance use. |
| Perceived easy availability was independently associated with recent substance use | aOR=3.22, 95% CI 1.42–7.30; p=0.005 | Participants reporting easy availability had higher adjusted odds of past-30-day substance use. |
| Social and environmental exposures interacted | aOR=1.51, 95% CI 1.01–2.27; p=0.047 | The association between social exposure and recent use differed according to perceived community availability. |
| Supply-side law enforcement was the most commonly proposed intervention | 124/285 (43.5%) | Restriction of local substance supply was the most frequently expressed community-proposed strategy. |
| Fatalism or resistance was relatively uncommon | 17/285 (6.0%) | A minority of respondents expressed scepticism about the possibility of reducing substance use. |

Qualitative findings showed that supply side law enforcement was the most frequently proposed community intervention (43.5%), while 6.0% of participants expressed fatalism or resistance regarding the possibility of reducing substance use.

## Discussion

This mixed-methods investigation demonstrates that adolescent and youth substance use in metropolitan Lagos is best understood as a product of interacting community environments, peer networks, and individual positions within those systems. By integrating multivariable quantitative modeling with youth-generated qualitative data, this study advances beyond descriptive epidemiology to elucidate the social ecology through which substances are accessed, normalized, and sustained. The most salient finding, that perceived community availability was independently associated with recent use and significantly modified the association between social exposure and recent substance use, supports a shift from exclusively individual-focused prevention toward place-based and socially embedded intervention strategies. The strong and independent association between perceived ease of access and past-30-day substance use is consistent with the availability hypothesis, which posits that reduced physical and social barriers to acquisition increase consumption at the population level. While this framework has been widely studied for alcohol and tobacco, emerging evidence suggests similar environmental influences across a broader range of substance use behaviors. A systematic review found that greater geographic availability of alcohol outlets was associated with higher adolescent drinking prevalence and riskier patterns of alcohol use.¹□ In parallel, research in low- and middle-income country (LMIC) contexts shows that environmental stressors and neighborhood structural conditions are important correlates of youth substance use. Broad syntheses of young people in sub-Saharan Africa highlight the substantial prevalence of alcohol, tobacco, khat, and other drug use and underscore the structural and community determinants in challenging urban contexts.²³ The present study extends this literature by showing that perceived easy availability remained strongly associated with past-30-day substance use (aOR = 3.22, 95% CI 1.42–7.30; p = 0.005) after adjustment for social exposure, demographic characteristics, and geographical study area fixed effects. This persistence suggests that perceived community availability may represent a distinct contextual exposure rather than merely reflecting social activity or individual risk. Ecological and longitudinal research similarly supports the proposition that neighborhood and built environment factors can independently influence adolescent substance involvement.²

The geospatial distributions presented for Yaba provide a granular articulation of how the built environment may structure substance use risk through the spatial co-location of mobility, commerce, and social congregation.¹ A higher past-30-day prevalence was observed in Makoko (34.5%), followed by Iwaya (23.6%) and Onike (23.4%), while a lower prevalence was observed in Alagomeji (19.7%) and Abule Oja (12.8%). Although these differences were not statistically significant across the study areas (χ²[4] = 7.46, p = 0.114), the mapped distributions remain informative for understanding how local environmental characteristics may intersect with patterns of recent substance use. Makoko, in particular, combined the highest past-30-day prevalence with the highest perceived availability (84.5%), mean Social Exposure Score (1.50), and prevalence of known substance-use locations (89.7%). These characteristics are consistent with the concept of high-access microenvironments, in which substances may become embedded within everyday transactional and social spaces and circulate through routine economic and interpersonal exchanges. ² ⁻² In contrast, Abule Oja, which recorded the lowest past-30-day prevalence (12.8%), also had the lowest mean Social Exposure (0.96) and perceived easy availability (57.4%). Iwaya occupied an intermediate position, with a past-30-day prevalence of 23.6%, a mean Social Exposure Score of 1.32, and a perceived easy availability of 76.4%. These patterns suggest that substance-use risk may not align linearly with deprivation, infrastructure, or any other single element of the urban environment. Rather, it may emerge from the intersection of social exposure, perceived availability, mobility, commercial activity, and local congregation within specific urban settings.¹□ Therefore, the broader spatial pattern is more consistent with a localized risk ecology than with a simple high-versus-low deprivation gradient.

The proximity of several study areas to major educational, transport, and commercial nodes may further contribute to these localized configurations by increasing population turnover, peer contact, and opportunities for substance acquisition.³ However, the present findings should be interpreted cautiously because the study did not directly quantify land-use density, walkability, outlet density, or distance to transportation and educational facilities. Therefore, the maps provide a contextual spatial interpretation rather than evidence that these built environment features independently caused the observed differences. Taken together, the findings support the view that in rapidly urbanizing LMIC settings, substance-use risk is shaped by the combined and potentially reinforcing effects of social and environmental conditions, rather than by isolated individual or neighborhood characteristics alone.

A key contribution of this study is the identification of a statistically significant interaction between social exposure and perceived availability. Specifically, the association between the Social Exposure Score and past-30-day substance use was stronger among participants who reported that substances were easily available in their communities. In the moderated model, the interaction term was statistically significant (aOR = 1.51, 95% CI 1.01–2.27; p = 0.047), while the conditional marginal effect of a one-unit increase in Social Exposure Score was +0.08 when easy availability was not reported and +0.18 when it was reported. This finding aligns with socio-ecological models in which environmental opportunity structures facilitate the translation of social norms and pressure into behavior. Systematic reviews and meta-analyses confirm that peer connectedness is a robust correlate of substance use among adolescents, with social networks and perceived peer norms being significantly associated with higher use across substances.² Network research further demonstrates that variations in peer group composition and structure are associated not only with substance-use behavior but also with broader mental health outcomes, underscoring the embeddedness of behavioral risks within social systems. Cross-national longitudinal evidence also suggests that peer substance use and communication patterns prospectively predict changes in adolescent alcohol and cannabis involvement over time.²□ The interaction observed here reinforces socio-ecological conceptions proposing that permissive environments may amplify the influence of social exposure by reducing barriers to substance access. Although many studies have examined peer effects independently, fewer have evaluated their interactions with environmental opportunity structures, particularly in urban LMIC settings.

The convergence of the quantitative findings with youth-articulated solutions further strengthens the mixed-methods interpretation. Supply side law enforcement was the most frequently proposed intervention theme, identified by 43.5% of participants, closely corresponding with the strong quantitative association between perceived easy availability and past-30-day substance use. This convergence suggests that participants recognized substance availability as an important community-level concern, although their recommendations should not be interpreted as evidence that enforcement is effective independently. Community-based prevention approaches have reduced adolescent substance use when environmental and social conditions are addressed concurrently.² Importantly, the detected interaction cautions against the reliance on a single intervention domain. Where substances are perceived to be readily available, social exposure may be more strongly associated with recent use, supporting the rationale for combining environmental strategies with interventions addressing peer and family influences. School- and community-based prevention models integrating peer engagement, parental involvement, and measures addressing local access and policy have been identified as important components of effective substance use prevention approaches in LMIC settings.²

Youth emphasis on economic empowerment and employment opportunities points to the broader structural conditions that shape vulnerability to substance use. This aligns with evidence from Africa, indicating that adolescent substance use is associated with sociodemographic and community-level factors, including age, sex, and exposure to peers who use substances.³□ Systematic research in sub-Saharan Africa similarly highlights higher substance-use prevalence among males and older adolescents in many settings, while peer networks and normative exposure consistently emerge as important correlates of substance use.³¹ Structural vulnerability frameworks further suggest that socioeconomic disadvantage can restrict access to protective resources and increase exposure to risk-promoting environments, particularly in areas where informal economies are prominent and regulatory structures are fragmented. In the present study, economic and youth empowerment was identified by 13.3% of participants as a potential response to substance use, indicating that some young people perceive employment, skills development, and broader socioeconomic opportunities as relevant components of prevention. The identification of a small but notable group expressing fatalism or resistance toward prevention efforts (6.0%) is also relevant to implementation research. If not adequately addressed, such attitudes may constitute contextual barriers to intervention uptake or sustainability. Contemporary implementation frameworks emphasize the early identification of contextual barriers and the use of participatory strategies, co-design processes, and transparent communication to improve the acceptability and feasibility of public health programmes among young people.³²

This study is strengthened by its community-based mixed-methods design, which enabled simultaneous examination of epidemiological patterns and youth-articulated responses within a single urban context. Integrating multivariate quantitative analyses with qualitative findings enhances explanatory depth and contextual interpretation, moving beyond isolated risk-factor identification toward a more ecologically grounded understanding of adolescent and youth substance-use behavior. Another methodological strength was the a priori specification and formal testing of the interaction between social exposure and perceived community availability, allowing for the assessment of whether the association between social exposure and recent substance use differed according to the perceived accessibility of substances. The statistically significant interaction observed in this study provides additional insight into how social and environmental exposures may operate jointly rather than independently of each other.

This study has several limitations that warrant careful consideration. The cross-sectional design precludes the determination of temporal sequence and limits causal inference, including the possibility that participants who use substances may perceive community availability differently from those who do not. Reliance on self-reported behaviors may have introduced recall or social desirability bias, although efforts were made to enhance privacy and use neutral question framing during the data collection. The use of selected geographical study areas within a single metropolitan setting also limits generalizability to other urban, peri-urban, or rural contexts with different social, cultural, regulatory, and substance market characteristics. In addition, environmental measures were based primarily on participant perceptions rather than objective measures of outlet density, geographic accessibility, or direct observation of substance availability. Although multivariable adjustment and interaction modeling accounted for several measured characteristics, residual and unmeasured confounding factors could not be excluded.

Furthermore, although the geospatial presentation provides useful contextual visualization, the study was not designed as a formal spatial epidemiological analysis and did not directly quantify land-use density, walkability, proximity to transport nodes, or substance-selling outlets.

Future studies should employ longitudinal designs to clarify the temporal ordering of community availability, social exposure, substance use initiation, and subsequent patterns of use. Geospatial methodologies incorporating systematically mapped substance outlets, objectively measured neighborhood characteristics, and appropriately protected location data could further examine the relationship between perceived and objectively measured availability. Implementation studies are also needed to evaluate community-informed multilevel strategies that combine environmental, interpersonal, family, and treatment-oriented components, particularly in rapidly urbanizing LMIC. Finally, the expanding role of digital and social media environments in shaping youth peer networks, substance-related norms, and potential access pathways warrants focused investigation alongside conventional neighborhood and community exposures.

## Conclusion

Adolescent and youth substance use in urban Lagos is embedded within the interaction of perceived community availability, social exposure, and broader contextual conditions. The findings indicate that perceived availability was independently associated with past-30-day substance use, and that the association between social exposure and recent use was stronger where substances were perceived to be readily available. These patterns underscore the need for integrated multilevel prevention strategies that address environmental access, strengthen family, peer, and community-level protective factors, and respond to wider socioeconomic vulnerabilities. By combining quantitative epidemiological evidence with youth-articulated perspectives on prevention, this study provides a contextually grounded basis for developing and evaluating locally appropriate substance-use prevention approaches in rapidly urbanizing settings in the future.

## Data Availability

De-identified datasets are available from the corresponding author upon reasonable request, in line with the data-sharing policy of this journal.

## Ethics Approval and Consent to Participate

Ethical approval was obtained from the Health Research Ethics Committee of the Lagos State Health Service Commission. The study was conducted in accordance with the Declaration of Helsinki and the Good Clinical Practice standards. All participants provided written informed consent before participation.

## Clinical trial number

Not Applicable

## Consent for Publication

Not applicable; no identifying individual data were reported.

## Availability of Data and Materials

De-identified datasets are available from the corresponding author upon reasonable request, in line with *the* data-sharing policy of this journal.

## Authors’ Contributions

All authors meet the ICMJE criteria for authorship and are accountable for the accuracy and integrity The contributions of this study are as follows:

- **AO, IAA, OO, OVO, AIO**: Study conceptualization, methodology design, data analysis, literature synthesis, manuscript drafting, manuscript editing, and manuscript revision.
- **AO, IAA, OO, OVO, AIO**: Data collection coordination, Data cleaning, statistical interpretation, and critical editing.
- **AO, IAA, OO, OVO, AIO**: Supervision of fieldwork and ethical compliance.

All authors have reviewed and approved the final manuscript and consented to its submission.

## Acknowledgements

We are grateful to the people who participated in this study. We also thank the research assistants and field staff for their dedication.

